# Virus detection by CRISPR-Cas9-mediated strand displacement in a lateral flow assay

**DOI:** 10.1101/2025.01.27.25320727

**Authors:** Roser Montagud-Martínez, Rosa Márquez-Costa, Raúl Ruiz, Adrià Martínez-Aviñó, Rafael Ballesteros-Garrido, David Navarro, Pilar Campins-Falcó, Guillermo Rodrigo

**Author notes:** Equal contribution (Roser Montagud-Martínez, Rosa Márquez-Costa).

## Abstract

In public health emergencies or in resource-constrained settings, laboratory-based diagnostic methods, such as RT-qPCR, need to be complemented with accurate, rapid, and accessible approaches to increase testing capacity, as this will translate into better outcomes in disease prevention and management. Here, we develop an original nucleic acid detection platform by leveraging the CRISPR-Cas9 and lateral flow immunochromatography technologies. In combination with an isothermal amplification that runs with a biotinylated primer, the system exploits the interaction between the CRISPR-Cas9 R-loop formed upon targeting a specific nucleic acid and a fluorescein-labelled probe to generate a visual readout on a lateral flow device. Our method enables rapid, sensitive detection of nucleic acids, achieving a limit of 1-10 copies/μL in 1 h at low temperature. We validated the efficacy of the method using clinical samples of patients infected with SARS-CoV-2. Compared to other assays, it operates with more accessible molecular elements and showcases a robust signal-to-noise ratio. Moreover, multiplexed detection was demonstrated using primers labeled with biotin and digoxigenin, achieving simultaneous identification of target genes on lateral flow devices with two test lines. We successfully detected SARS-CoV-2 and Influenza A (H1N1) in spiked samples, highlighting the potential of the method for multiplexed diagnostics of respiratory viruses. All in all, this represents a versatile and manageable platform for point-of-care testing, thereby supporting better patient outcomes and enhanced pandemic preparedness.

## INTRODUCTION

Nucleic acid detection has become a standard approach for precise disease diagnosis thanks to the rapid advancement of genetic and genomic sequencing technologies [1]. For infectious diseases, accurately identifying the causative pathogen is essential for implementing the correct treatment and containment strategies, requiring in most cases multiplexed reactions [2]. In the case of non-transmissible diseases, detecting key genetic variations and biomarkers can enable early intervention before symptoms appear and the condition worsens [3]. Apart from clinical diagnostics, nucleic acid detection with high specificity and sensitivity is vital for applications in epidemiological surveillance, agricultural biosecurity, environmental monitoring, and biodefense.

Polymerase chain reaction (PCR) methods are the cornerstone of diagnostics across various fields [4]. However, the coronavirus disease 2019 (COVID-19) pandemic [5] has underscored the need for diagnostic tools beyond PCR; ones that do not require expensive equipment or specialized training, hence facilitating large-scale screening programs. According to the World Health Organization, these new tools should be affordable, sensitive, specific, user-friendly, rapid, equipment-free, and deliverable (ASSURED) [6]. These characteristics make them ideal for point-of-care (POC) testing and use in low-resource settings. Notably, by increasing testing capacity through a combination of complementary methods, better outcomes in disease prevention and community-level mitigation could be achieved [7].

In recent times, systems based on clustered regularly interspaced short palindromic repeats (CRISPR) have found innovative applications in the realm of specific and sensitive nucleic acid detection [8]. These ribonucleoprotein-based systems are archetype because of their programmable sequence specificity, diverse binding and cleavage mechanisms, and compatibility with advanced nanobiotechnological developments. Pioneering work disclosed the *trans*-cleavage activity of the nucleases Cas12a and Cas13a (on single-stranded DNA and RNA, respectively) [9,10], which was exploited to produce a suitable readout upon target recognition in combination with an isothermal pre-amplification step. Recombinase polymerase amplification (RPA) [11] is particularly favored for its simplicity in design and execution, though other amplification methods could also be employed [12]. The resulting CRISPR-based approach allows detections with single nucleotide specificity and attomolar sensitivity, and even the use of lateral flow immunochromatographic assays (LFAs) for their resolution. However, in these applications with commercially available strips the control line is used as the test line and *vice versa*, which greatly reduces the sensitivity of the technique and favors the occurrence of false positives [13].

The CRISPR-Cas9 system can also be repurposed for nucleic acid detection. CASLFA (CRISPR-Cas9-mediated lateral flow nucleic acid assay) [14], FELUDA (*Francisella novicida* Cas9 editor linked uniform detection assay) [15], Bio-SCAN (biotin-coupled specific CRISPR-based assay for nucleic acid detection) [16], and Vigilant (VirD2-dCas9 guided and LFA-coupled nucleic acid test) [17] are methods that have been developed to exploit LFA kits, relying on RPA and CRISPR-Cas9. These techniques depend on the formation of a suitable CRISPR-dCas9-DNA complex (with dCas9 being a catalytically inactive nuclease) carrying biotin and fluorescein labels. In particular, CASLFA utilizes a biotinylated primer and a gold nanoparticle-linked oligonucleotide probe, FELUDA uses a biotinylated primer and fluorescein-labelled single guide RNA (sgRNA), Bio-SCAN employs a fluorescein-labelled primer and a biotinylated dCas9, and Vigilant combines a biotinylated primer with a dCas9-relaxase fusion attached to a fluorescein-labelled oligonucleotide.

In the aforementioned diagnostic schemes, the test line, which is more sensitive than the control line, provides the readout. They have demonstrated limits of detection ranging from attomolar to femtomolar within approximately one hour and are capable of discriminating small genetic variations. Of note, they have been effectively applied to detect pathogens such as severe acute respiratory syndrome coronavirus 2 (SARS-CoV-2), African swine fever virus (ASFV), and *Listeria monocytogenes*. However, these methods require the modification of either the sgRNA or the nuclease, which prevents a widespread and cost-effective application. In addition, despite some efforts have been done with CASLFA [18], the multiplexed detection of different species in the same reaction with the CRISPR-Cas9 system to then be resolved in an LFA needs to be investigated (*e*.*g*., to discriminate infections produced by different viruses).

Recently, we developed a novel nucleic acid detection method based on CRISPR-Cas9-mediated strand displacement, which we called COLUMBO (CRISPR-Cas9 R-loop usage for molecular beacon opening) [19]. This method allowed a multiplexed detection in a single tube and demonstrated biocomputing capabilities [20], thereby complementing the existing CRISPR-based diagnostic techniques. Nonetheless, it relies on the generation of a fluorescence signal upon nucleic acid recognition, which precludes its pervasive application in POC settings. In this work, we report the modification of COLUMBO aimed to exploit LFAs to resolve the detection. To this end, we employed biotin- and digoxigenin-labelled primers for the pre-amplification step and fluorescein-labelled probes for the CRISPR-Cas9-mediated strand displacement reactions. Our results show that a rapid and streamline procedure implemented with attainable reagents allows the multiplexed detection of different species and displays ASSURED diagnostic potential.

## RESULTS AND DISCUSSION

### Molecular mechanism and functional characterization

We previously showed that a CRISPR-Cas9 ribonucleoprotein appropriately programmed to target a given DNA molecule, typically obtained by a pre-amplification step, generates an R-loop that can be exploited to interact with a molecular beacon supplied in *trans* by means of a toehold-mediated strand displacement reaction, thereby producing a fluorescence readout [19]. Yet, the detection could also be resolved in an LFA with subtle adaptations. In typical commercial paper-based devices, gold nanoparticles functionalized with anti-fluorescein antibodies are used to detect the presence of a specific analyte by observing a color change in a test line that is coated with Streptavidin [13]. Here, we envisioned the use of a biotin-labelled primer to generate DNA amplicons that can be recognized by Streptavidin, and the use of a fluorescein-labelled probe that can interact with the displaced strand of the R-loop in the protospacer adjacent motif (PAM)-proximal region to be recognized by the anti-fluorescein antibody. Thus, the formation of the quaternary complex upon DNA targeting by the CRISPR-Cas9 ribonucleoprotein and subsequent interaction with the probe would lead to a color change in the test line (**Fig. 1**). The intensity of such a band can then be quantified for an accurate result. Nothing prevents using a fluorescein-labelled primer and a biotin-labelled probe to obtain a similar visual output. However, to implement a multiplexed detection, it would be convenient to have biotin- and digoxigenin-labelled primers together with fluorescein-labelled probes to exploit commercial LFA strips.

**Fig. 1:** Schematic overview of iCOLUMBO. The method can be applied to detect any nucleic acid (DNA or RNA), including viral genomes. The iCOLUMBO assay can be completed in less than 1 h from the collected patient sample to the LFA strip with the result. On the top, illustration of the procedure and molecular mechanism of iCOLUMBO. It uses RT-RPA to implement a pre-amplification step with a biotin-labelled primer (recognized by Streptavidin) and then a programmed sgRNA and a fluorescein-labelled probe (recognized by the anti-fluorescein antibody) for the specific CRISPR-Cas9-based detection of the resulting DNA amplicon. On the bottom, illustrative scheme of the functioning of the LFA strip. The formation of the quaternary complex upon specific DNA detection by the CRISPR-Cas9 ribonucleoprotein and subsequent probe binding leads to a color change in the test line. B7, biotin. FAM, fluorescein.

To prove the functionality of the system, we considered the RNA genome of SARS-CoV-2 as the nucleic acid of interest. Using DNA amplicons from the N and E genes generated with custom primers by reverse transcription PCR (RT-PCR), we showed good detection resolution in the LFA with ribonucleoproteins appropriately programmed (**Fig. 2A**). In absence of sgRNA or Cas9, the test line did not reveal a noticeable signal, stressing that the detection was mediated by the CRISPR-Cas9 complex. A quantification of the band intensity in the test line for increasing concentrations of input DNA revealed a monotonous response and a sensitivity of 5-10 nM (**Fig. 2B**; see replicates in **Fig. S1**). Using dilutions of the RNA genome and RT-RPA for the pre-amplification step, we found a limit of detection for the whole technique of 1-10 copies/μL (**Fig. S2**). Importantly, this limit of detection is comparable to that of quantitative RT-PCR (RT-qPCR). In addition, we examined the impact of the catalytic activity of Cas9 on the performance of the assay. We used four different versions of Cas9 to this end, *viz*., the wild-type nuclease (from *Streptococcus pyogenes*), the Cas9 H840A nickase (Cas9n), which only cleaves the non-targeted strand, the catalytically dead Cas9 protein (dCas9), which does not produce any cleavage, and an engineered version of Cas9 with reduced off-target effects (Cas9HF). All nucleases were able to perform the proper detection (**Fig. 2C**). However, we observed that the use of Cas9n produced less visual signal in the test line. These results demonstrate the suitability of this new method, dubbed iCOLUMBO (immunochromatographic COLUMBO), to perform POC testing.

**Fig. 2:** Detection of DNA with iCOLUMBO. A) Image of representative LFA strips in the detection of the DNA amplicons from the N and E genes of SARS-CoV-2. i) N gene amplicon. ii) E gene amplicon. B) Image of representative LFA strips (right) and quantified intensity of the test line band (left) for increasing concentrations of DNA (from 0 to 40 nM).C) Image of representative LFA strips in the detection of DNA for different versions of Cas9. Represented data in the bar plots correspond to means ± standard deviations (*n* = 3). Statistical significance assessed by the Welch’s *t*-test, *P* < 0.05. *Statistically significant change, ^ns^non-significant change (comparisons against 0). AU, arbitrary units.

### Mutant discrimination, multiplexed detection, and comparative performance

The ability to detect mutations with a CRISPR-based system may be important to discriminate between strains or variants of an infectious agent, without requiring sequencing, thereby providing rapid and cost-effective insights about the transmission dynamics [21]. We introduced artificial mutations in the E gene amplicon and assessed the ability of iCOLUMBO to detect them. The system was responsive to a mutation in the PAM, but not to a mutation in the protospacer (in the PAM-proximal region; **Fig. 3A**). The use of Cas9HF neither resulted effective to detect the mutation in the protospacer, despite we previously found that this engineered nuclease was instrumental to do so with COLUMBO [19]. Arguably, even a slight proportion of quaternary complexes formed are relevant because of the higher sensitivity of the LFA. With Bio-SCAN, for instance, the mutations that were detected also affected the PAM or were deletions of more than one nucleotide [16]. Further work is required to engineer systems suitable to detect arbitrary mutations. In addition, the simultaneous detection of two different regions of the pathogen genome may be instrumental to maximize the accuracy of the detection by surmounting scenarios at the edge of the signal-to-noise limit. For that, we employed LFA strips with two test lines, one coated with Streptavidin and another with an anti-digoxigenin antibody. We performed the multiplexed detection of the N and E gene amplicons, showing a proper detection resolution in the LFA (**Fig. 3B**; see also a movie of the procedure in **WEO S1**).

**Fig. 3:** Detection of mutations and multiple genes with iCOLUMBO. A) Image of representative LFA strips (right) and quantified intensity of the test line band (left) in the detection of DNA (E gene of SARS-CoV-2) with different mutations, which are shown in red in the inset. B) Image of representative two-test-line LFA strips, together with an illustrative scheme of the functioning, in the multiplexed detection of DNA (N and E genes of SARS-CoV-2). C) Comparative performance assessment. Scheme of the CRISPR-based detection and image of LFA strips in the detection of DNA (N gene of SARS-CoV-2). i) CRISPR-Cas9. ii) CRISPR-Cas12a. Represented data in the bar plot correspond to means ± standard deviations (*n* = 3). Statistical significance assessed by the Welch’s *t*-test, *P* < 0.05. *Statistically significant change, ^ns^non-significant change (comparisons against WT). B7, biotin. DIG, digoxigenin. FAM, fluorescein. AU, arbitrary units.

To realize about the nucleic acid detection potential of iCOLUMBO, we aimed to compare its performance with that of DETECTR (DNA endonuclease-targeted CRISPR *trans* reporter) [9]. This latter method is based on the CRISPR-Cas12a system and has been widely applied in recent times, such as to detect SARS-CoV-2 [22] or monkeypox virus [23]. To perform the CRISPR-Cas12a reaction here, we used the nuclease from *Lachnospiraceae bacterium* and a probe labelled with fluorescein and biotin. We focused on the detection of the SARS-CoV-2 N gene amplicon. With CRISPR-Cas9, the test line was almost clean to the naked eye in absence of target DNA and the control line was always colored. By contrast, with CRISPR-Cas12a, a faint band was observed in the test line in absence of target DNA and the band in the control line changed according to the input (**Fig. 3C**). This stresses the distinctive usage of the commercial LFA strips with both systems. It is important to note that while the *trans*-cleavage activity of Cas12a serves to obtain a higher dynamic range in a fluorescence-based assay, the visual signal obtained in an LFA upon Cas9-mediated assembly is more differential (**Fig. S3**).

Notwithstanding, it has been reported a *trans*-cleavage activity of Cas9 when using a poly(T) probe [24]. This activity appears to be increased if the ribonucleoprotein is formed with a CRISPR RNA (crRNA) and a *trans*-activating crRNA (tracrRNA), *i*.*e*., the natural form. Using a crRNA targeting the E gene amplicon and the universal tracrRNA, we did not find any degradation effect on the specific fluorescein-labelled probe (**Fig. S4**), suggesting that the eventual collateral activity of Cas9 is irrelevant within the iCOLUMBO setup.

### Validation with clinical samples

To illustrate the applicability of the technology, we collected clinical samples from patients showing symptoms compatible with COVID-19 in order to perform virus detection assays. Initially, we confirmed the presence of SARS-CoV-2 in the samples by RT-qPCR using the Centers for Disease Control and Prevention (CDC) N1 primers (**Fig. S5**). Infected patient samples showed cycle threshold (C_T_) values lower than 30. Subsequently, we ran CRISPR-Cas9-based reactions. We found that the iCOLUMBO assay, both targeting the N and E genes, gave marked differential readouts in the LFA strips that were useful to discriminate the presence of the virus (**Fig. 4A**). We used custom primers targeting the N and E genes of SARS-CoV-2 and RT-RPA (without RNA extraction) for the pre-amplification step.

**Fig. 4:** Detection of SARS-CoV-2 in clinical samples with iCOLUMBO. A) Image of representative LFA strips (right) and quantified intensity of the test line band (left) in the detection of the RNA genome of SARS-CoV-2. i) N gene amplicon. ii) E gene amplicon (pre-amplification by RT-RPA). B) Image of representative two-test-line LFA strips (bottom) and quantified intensities of the test line bands (top) in the multiplexed detection of SARS-CoV-2. Represented data in the bar plots correspond to means ± standard deviations (*n* = 3). Statistical significance assessed by the Welch’s *t*-test, *P* < 0.05. *Statistically significant change, ^ns^non-significant change (comparisons against P2). B7, biotin. DIG, digoxigenin. FAM, fluorescein. AU, arbitrary units.

In addition, we performed the multiplexed detection of the N and E genes, also obtaining marked differential readouts in the LFA strips with two test lines (**Fig. 4B**). In this case, RT-RPA was regularly performed with two pairs of primers, one of them targeting the N gene labelled with biotin and another targeting the E gene labelled with digoxigenin. This set of primers showed good compatibility for the multiplexed amplification. Next, CRISPR-Cas9-based reactions with two different ribonucleoproteins and probes were carried out. Compared to the single detection, we noticed more variability between replicates in the results. We also observed that the system performance could be fine-tuned by optimizing the relative concentrations of primers and probes used in the reactions.

These results demonstrate the potential of iCOLUMBO to perform *in vitro* clinical diagnostics in a simple and effective way. A number of considerations can be drawn. First, RNA extraction and concentration (*e*.*g*., through magnetic beads) are considered as rate-limiting steps and potential sources for contamination. Our approach does not involve such steps and allows working directly with the collected sample. Still, further work is required to combine the RT-RPA and CRISPR-Cas9 reactions to end with a one-pot detection procedure. Second, reactions are implemented with highly attainable molecular and instrumental elements. Specific nucleic acids can be redesigned and easily resynthesized. In the clinic, a computational method could be used to automate such a design given the full sequence(s) of interest. Third, iCOLUMBO has the potential to target a wide range of infectious agents, including viruses and bacteria. Besides, the assay could be exploited to recognize nucleic acids beyond pathogen genomes, such as specific genetic markers associated with non-communicable diseases (*e*.*g*., cancer), providing a powerful tool for early detection and management.

### Multiplexed detection of clinically relevant respiratory viruses

Infections provoked by different respiratory viruses usually cause similar symptoms. Therefore, it is important to have methods specific enough to identify the causing agent and adopt the appropriate treatment. These viruses can even coinfect the host organism [25]. Mixed infections are less frequent but lead to more severe outcomes for the patient, especially in children. In this regard, we assessed the ability to detect simultaneously two different viruses in the sample with iCOLUMBO. We focused on SARS-CoV-2 and Influenza A (H1N1). While SARS-CoV-2 has caused the recent COVID-19 pandemic, the evolution and transmission of Influenza has increased with time, so there is also an elevated risk for public health associated with this virus [26]. In addition, correctly identifying infections caused by viruses rather than bacteria in the respiratory tract is crucial for mitigating the overuse of antibiotics. Viral infections, such as those caused by SARS-CoV-2 or Influenza, do not respond to these compounds. Yet, misusing antibiotics for viral infections contributes to the emergence and evolution of multidrug resistant bacteria, which is a growing global health challenge [27].

Thus, artificial samples spiked with the RNA genomes of these viruses in a combinatorial way were prepared. A concentration of ∼10^3^ copies/μL was considered, as it is comparable to those found in clinical samples. We designed primers to amplify the M gene of Influenza A (H1N1) and a new sgRNA and probe to detect the resulting DNA amplicon with CRISPR-Cas9. One of these new primers was labelled with biotin to be used in combination with those targeting the E gene of SARS-CoV-2 (labelled with digoxigenin) in the multiplexed RT-RPA and subsequent detection step. Multiplexed amplifications are always critical and may require screening various sets of primers; the primers here considered showed adequate compatibility (**Fig. S6**). In first place, we verified that the single detection of Influenza A (H1N1) was possible with the designed elements (**Fig. S7**). Then, we found that iCOLUMBO allowed the multiplexed detection of the case study respiratory viruses employing the LFA strips with two test lines (**Fig. 5**). The method distinguished between samples containing either SARS-CoV-2, Influenza A (H1N1), or both, without noteworthy cross-reactivity. Yet, we noticed that the band intensity in the first test line was lower, suggesting that the designed elements to detect Influenza A (H1N1) were less efficient than those to detect SARS-CoV-2, especially the amplification primers. These findings demonstrate that the challenge of coinfection diagnostics can be addressed with CRISPR systems. Ultimately, our development is aligned with ensuring better patient care and strengthening pandemic preparedness efforts.

**Fig. 5:** Multiplexed detection of SARS-CoV-2 and Influenza A (H1N1) with iCOLUMBO. Image of representative two-test-line LFA strips (right) and quantified intensities of the test line bands (left) in the multiplexed detection of the RNA genomes of SARS-CoV-2 and Influenza A (H1N1) (pre-amplification by RT-RPA). Represented data in the bar plot correspond to means ± standard deviations (*n* = 3). Statistical significance assessed by the Welch’s *t*-test, *P* < 0.05. *Statistically significant change, ^ns^non-significant change (comparisons against ∅). SARS2, SARS-CoV-2. InfA, Influenza A (H1N1). B7, biotin. DIG, digoxigenin. AU, arbitrary units.

## CONCLUSION

This study sought to develop a nucleic acid detection method applicable to virus detection that effectively balanced specificity, sensitivity, and POC potential. To this end, we relied on the CRISPR-Cas9 system for the detection, with a pre-amplification step of RT-RPA, and the LFA technology to produce a visual output. The proposed method does not rely on *trans*-cleavage but on logic nucleic acid intermolecular interactions, and it uses commercial LFA strips for a simple and cost-effective detection. The iCOLUMBO assay is completed in less than 1 h with high specificity and sensitivity, hence providing rapid, visual, and precise results. It does not require an initial step of RNA extraction or concentration, shows an attomolar limit of detection (*i*.*e*., <100 copies/μL in the collected sample), and allows a multiplexed detection of different nucleic acid species. It operates at a low constant temperature (37-42 °C), eliminating the need for temperature cycling. By employing CRISPR-Cas9 complexes for posterior recognition, the method effectively reduces the false positives generated by spurious amplifications, then enhancing specificity, in addition to provide a way to identify mutations, especially those that affect a PAM (with the aim to genotype a pathogen). Importantly, the evaluation of various clinical nasopharyngeal swab samples revealed that iCOLUMBO displayed the required specificity and sensitivity to obtain reliable diagnoses in the clinic, hence being a potential complement to the gold-standard RT-qPCR technique. It was applied to detect the RNA genomes of SARS-CoV-2 and Influenza A (H1N1). Furthermore, this assay may be accessible in low-income countries to monitor and control the spread of infectious diseases, which cause millions of deaths every year (especially those that affect the respiratory tract) [28]. With an in-house production system of purified enzymes and LFA strips, the unit cost of the assay may be drastically reduced and be competitive in the market. In sum, this advance for nucleic acid detection by leveraging the CRISPR-Cas9 system may significantly contribute towards the effective deployment of ASSURED diagnostic techniques in POC settings. We envision exciting applications of iCOLUMBO in the close future.

## METHODS

### Clinical samples

Nasopharyngeal swab samples from patients infected with SARS-CoV-2 and non-infected patients were gathered in the Clinic University Hospital of Valencia (Spain). Samples were treated with proteinase K followed by a heat shock (5 min at 60 °C) before proceeding. No RNA extraction was performed. The ethics committee of the Clinic University Hospital approved this study (order #2020/221).

### Spiked samples

Samples of total RNA from human embryonic kidney (HEK293T) cells at 5 ng/μL, extracted using RNAzol RT (Merck), were spiked with the complete and purified RNA genomes of SARS-CoV-2 and Influenza A (H1N1) (Vircell) in a combinatorial way at the final concentration of 5·10^3^ copies/μL each. These samples were used to perform the multiplexed detection of different respiratory viruses.

### Purified DNA amplicons

From the complete RNA genome of SARS-CoV-2 (Vircell), RT-PCRs were performed to generate suitable DNA amplicons of the N and E genes. The TaqPath 1-step RT-qPCR master mix, CG (Applied) was used, with 52 copies/μL RNA genome and 250 nM custom primers. The reverse primer was labelled with biotin or digoxigenin in its 5’ end. The protocol consisted of an initial step at 25 °C for 2 min for uracil-N-glycosylation and 50 °C for 15 min for RT, then an inactivation step at 95 °C for 2 min, followed by 40 cycles of amplification at 95 °C for 15 s and 60 °C for 60 s. RT-PCR products were purified using the DNA clean and concentrator column (Zymo) and quantified in a spectrophotometer (NanoDrop, Thermo). Besides, E gene amplicons with different single base mutations were chemically synthesized (IDT), then amplified by PCR to label with biotin.

### CRISPR elements

Four versions of S. pyogene*s* Cas9 (IDT) were used: the wild-type nuclease (Cas9), the Cas9 H840A nickase (Cas9n), the catalytically dead Cas9 protein (dCas9) [29], and a high-fidelity nuclease with similar on-target potency to the wild-type but significantly reduced off-target effects (Cas9HF). The *L. bacterium* Cas12a (NEB) was used to implement the CRISPR-Cas12a reactions. In addition, sgRNAs were produced by *in vitro* transcription with the TranscriptAid T7 high yield transcription kit (Thermo) from DNA templates (sequences provided in **Table S1**). They were purified using the RNA clean and concentrator column (Zymo) and quantified in a spectrophotometer (NanoDrop, Thermo).

### Nucleic acid detection by RT-qPCR

The TaqPath 1-step RT-qPCR master mix, CG (Applied) was used. In a microplate (Applied), 2 μL clinical sample, 500 nM CDC N1 primers, 125 nM probe (SARS-CoV-2 RUO kit, IDT), and RT-qPCR mix were combined for a total volume of 20 μL. The microplate was loaded into a real-time PCR system (QuantStudio 3, Applied) for fluorescence measurement, and the following protocol was employed: an initial step of 25 °C for 2 min for uracil-N-glycosylation, then a step of 53 °C for 10 min for RT, 95 °C for 2 min for RT inactivation, followed by 40 cycles of 95 °C for 3 s for denaturation and 60 °C for 30 s for annealing and extension. Samples with C_T_ values lower than 40 were considered positive for SARS-CoV-2 infection.

### Nucleic acid amplification by RT-RPA

The TwistAmp basic kit (TwistDX) was used. 500 U RevertAid (Thermo), 50 U RNase inhibitor (Thermo), and 480 nM custom forward and reverse primers were added to 29.5 µL rehydration buffer. Primers targeting the N and E genes of SARS-CoV-2 and the M gene of Influenza A (H1N1) were used (sequences provided in **Table S1**). The reverse primer was labelled with biotin or digoxigenin in its 5’ end. In the case of multiplexed amplifications, 360 nM primers were used (unless otherwise specified). The TwistAmp basic reaction pellet was resuspended, then splitting into two the resulting volume. To start the reaction, 1 µL patient or spiked sample and 7 mM magnesium acetate were added. Reactions were incubated at 42 °C for 30 min in a thermomixer (Eppendorf).

### Nucleic acid detection with CRISPR-Cas9

CRISPR reactions were performed in 1x Tris/Acetate/EDTA (TAE) buffer pH 8.5 (Invitrogen), 0.05% Tween 20, and 12.5 mM MgCl_2_ at a final volume of 20 µL. In the case of DNA amplicon detection (from the E and N genes of SARS-CoV-2), 100 nM CRISPR-Cas9 ribonucleoprotein, previously assembled at room temperature for 30 min, was mixed with 40 nM amplified DNA (unless otherwise specified) and 100 nM ssDNA probe (labelled with fluorescein). For the SARS-CoV-2 RNA genome detection in clinical samples, 2 µL RT-RPA product was used rather than a definite DNA amount. For the multiplexed detection of the SARS-CoV-2 E and N genes, the probe interacting with the E amplicon was added at 50 nM, while the concentration of all other species remained unaltered. For the multiplexed detection of the SARS-CoV-2 and Influenza A (H1N1) RNA genomes in spiked samples, 1 µL RT-RPA product and 100 nM of each of the probes were used. Reactions were incubated at 37 °C for 20 min in a thermomixer (Eppendorf).

### Nucleic acid detection with CRISPR-Cas12a

Reactions were performed in NEBuffer 2.1 (10 mM Tris-HCl pH 7.9, 50 mM NaCl, 10 mM MgCl_2_, and 100 μg/mL bovine serum albumin; NEB). 50 nM CRISPR-Cas12a ribonucleoprotein, previously assembled at room temperature for 30 min, was mixed with 40 nM amplified DNA and 100 nM ssDNA probe (labelled with fluorescein and biotin) to make a total volume of 20 μL. Reactions were incubated at 37 °C for 20 min in a thermomixer (Eppendorf).

### Lateral flow assay

After completing the CRISPR reactions, 80 µL GenLine Dipstick buffer (Milenia) was carefully added for a 1:5 dilution. The LFA strip (HybriDetect, Milenia) was soaked into the reaction tube for 4 min and images were captured with a gel documentation system (Uvidoc HD6, Uvitec). Quantification of the band intensity in the test line was done with Fiji [30]. Moreover, to perform a quantitative characterization of the strip, different mixtures of fluorescein, biotin, and ssDNA probe labelled with both fluorescein and biotin were prepared, maintaining the total level of biotin at 0.1 or 25 nM. The band intensities in the test and control lines were monitored by measuring absorbance with a conventional spectrophotometer (UV-Vis diffuse reflectance spectroscopy; **Figs. S8, S9**).

### Gel electrophoresis

Nucleic acid amplifications by PCR or RPA were confirmed by agarose gel electrophoresis. For that, 2 µL amplified product was used. Samples were loaded on a 3% agarose gel prepared with 0.5x Tris/Borate/EDTA (TBE) buffer, which was run for 45 min at room temperature (110 V). Gels were stained using GreenSafe (NZYtech). The GeneRuler ultra-low range DNA ladder (10-300 bp, Thermo) was used as a marker.

## Data Availability

All data produced in the present study are available upon reasonable request to the authors

## ACKNOWLEDGEMENTS

Work supported by CRUE and Banco Santander (Fondo Supera Covid-19), CSIC PTI+ Global Health (NextGenerationEU/PRTR), Spanish Ministry of Science and Innovation and AEI/10.13039/501100011033 (PDC2022-133941-I00 and PRE2019-088531, co-financed by NextGenerationEU/PRTR), and Regional Government of Valencia (SEJI/2020/011, GVA-COVID19/2021/036, CIPROM/2022/21, and CIPROM/2023/46).

## SUPPORTING INFORMATION

Additional experimental results of nucleic acid detection (**Figs. S1-S9**), nucleotide sequences of the elements used in this work (**Table S1**), and a movie showing a representative multiplexed detection (**WEO S1**).

## NOTES

RMM, RMC, RR, and GR declare that they have filled a patent on the use of the CRISPR-Cas9 R-loop for molecular beacon opening.

